# A Nursing Philosophy of Women Empowerment Based on Self Efficacy toward Sexual and Reproductive Health

**DOI:** 10.1101/2022.10.03.22280632

**Authors:** Retnayu Pradanie, Moses Glorino Rumambo Pandin, Esti Yunitasari

## Abstract

**Introduction:** Sexual and reproductive health of women will affect their health in general at every stage of their lives. Empowerment of women is a strategic step to improving health status. Nurses have an essential role to empower women to improve their health. Women empowerment can be built by improving their self-efficacy. This paper aimed to provide a review of articles about the role of nurses and strategies to empower women in sexual and reproductive health based on self-efficacy.

**Method:** This paper is the result of a literature review. Data were accessed from the database of Scopus, Pubmed, and Ebscohost limited on publication in 2017-2022. Twelve journal articles were selected using Preferred Reporting Items for Systematic Reviews and Meta-Analyses (PRISMA).

**Results:** The results of the article review obtained information about the important role of women in the health sector, factors associated with sexual and reproductive health, nursing philosophy women health, women empowerment models for sexual and reproductive health, and strategies to increase women empowerment through self-efficacy enhancement.

**Conclusion:** An evidence-based practice approach found that women empowerment is a strategic step for health development. The women empowerment program still needs to be developed because it can increase their capacity in maintaining health.

## Introduction

Nursing is a form of health service, based on nursing knowledge and tips, in the form of comprehensive bio-psycho-socio/cultural and spiritual services, aimed at individuals, families, and communities, both sick and healthy, covering all life processes (Budiono, 2016). As a form of professional service, nursing as a profession is responsible for realizing the nation’s health development. A healthy and highly competitive nation is born from a generation that is healthy mentally, physically, spiritually, and socially. Women have enormous potential as a pillar of national development. This potential can be seen in women carrying out the functions of their reproductive system when they are pregnant and giving birth to children as the next generation of the nation. Women must stay healthy to be able to raise and educate children with full attention and affection to lead children to adult life. Healthy women will also be able to carry out emancipation in the fields of education, economy, politics, and socio-culture. Thus, maintaining women’s health is very important, not only for the health and wellbeing of women themselves but also for their families and society at large (Kementerian Pemberdayaan Perempuan dan Perlindungan Anak Republik Indonesia, 2021).

Women’s health is the main focus of health development in the world (United Nations, 2022). This is evidenced by the establishment of Sustainable Development Goals (SDGs) on the 3rd goal (good health and well-being) and 5th goal (gender equality). The concept of women’s health uses a women’s life cycle approach and health services are carried out from the fetus to the grave (from womb to tomb) or also known as the “Continuum of care women cycle” (Prijatini & Rahayu, 2016). This is because women’s health in childhood will affect women’s health as adults, during pregnancy, childbirth, postpartum, and continues until menopause.

Along with the times, women also face various challenges in the health sector. The World Health Organization (WHO) has set 6 priorities related to women’s health issues which include increasing gender equality, improving the position of women in the health workforce, preventing and overcoming violence against women, ensuring quality sexual and reproductive health, suppressing non-communicable diseases among women, and increasing women’s participation in science development (World Health Organization, 2021). Various studies have been carried out to explain the factors that affect women’s health and the efforts that can be made to improve it. However, until now the main obstacles to women’s health are socio-cultural factors, economy, education, and the health care system (World Health Organization, 2022).

Empowerment of women is a strategic step to improve health status (Belz, 2018). Women’s empowerment is the ability of women to make decisions from the various options provided according to their wishes and get results from those choices (Habib Sultan & Yahaya, 2020). Improving women’s empowerment can be done through socialization and education to increase knowledge and attitudes to create healthy life behaviors (Kafaei-Atrian et al., 2022). Good knowledge and attitudes will ultimately increase self-efficacy and it has an impact on healthy living behavior (Bandura, 2009). Nurses through their roles as caregivers and educators have a great contribution to women’s empowerment programs to improve health (Brownie et al., 2018). The model of women’s empowerment in terms of nursing is very important to continue to be developed. Through empowerment, women can live safely, adequately, and productively so that they can make a complete contribution to their families and society. This study aims to explain the potential for women’s empowerment based on self-efficacy on sexual and reproductive health.

## Method

This was a systematic review using data from the database of Scopus, Pubmed, and Ebscohost limited on publication in 2017-2022. The search strategy using keywords that was determined from the PICO framework included Population (women OR girls OR mothers), Interests/Intervention (women empowerment AND self-efficacy), Comparison (Nursing intervention), and Outcome (women’s health OR sexual reproductive health). All articles obtained were analyzed using a PRISMA flow chart to obtain relevant articles.

## Results

A total of 12 articles that matched specified keywords were obtained from Scopus (n=588), Pubmed (n=5154), and Ebscohost (n=26354). The entire article was then analyzed using a PRISMA flow chart as shown in Figure 1 and the results of the articles obtained are presented in table 1.

**Table 1.**
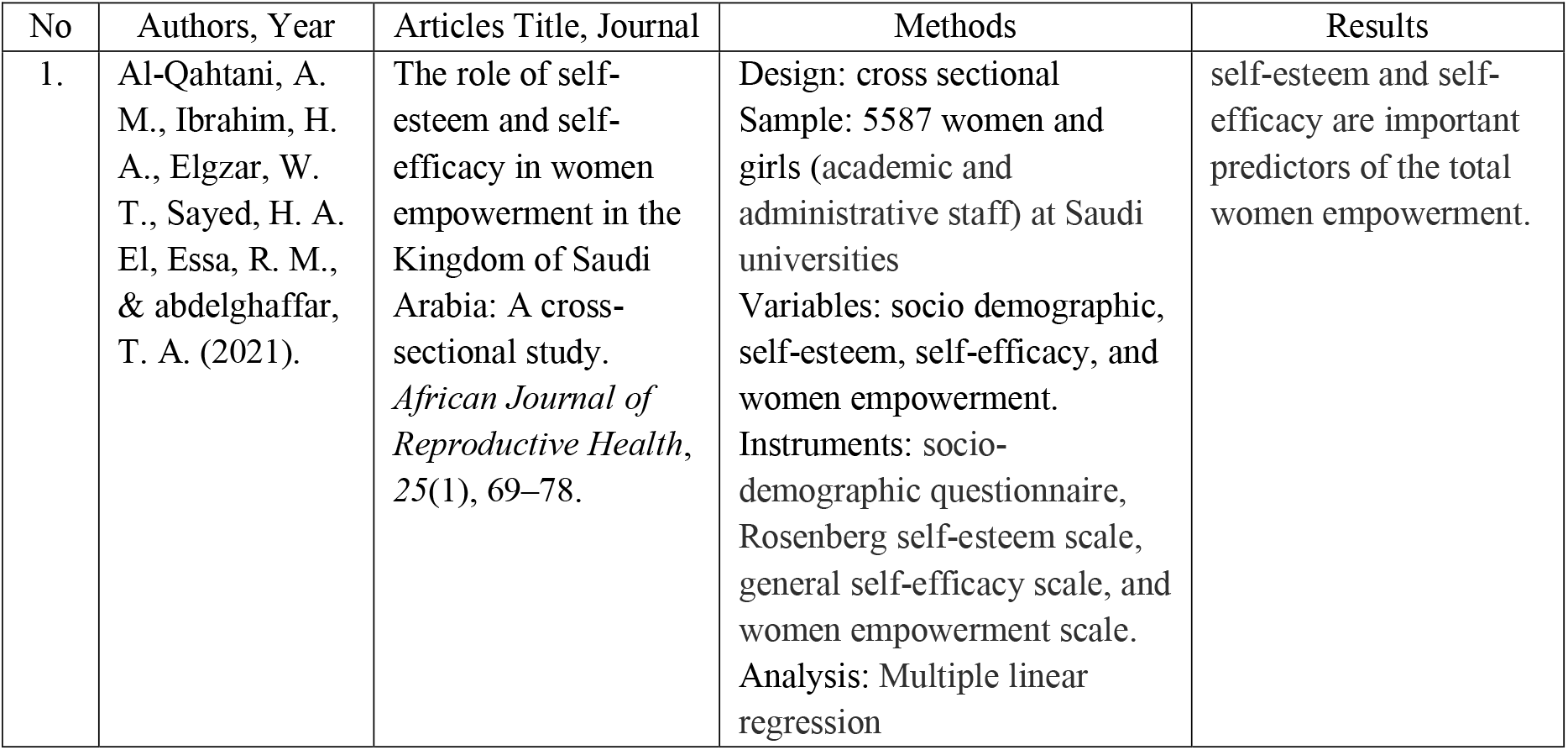
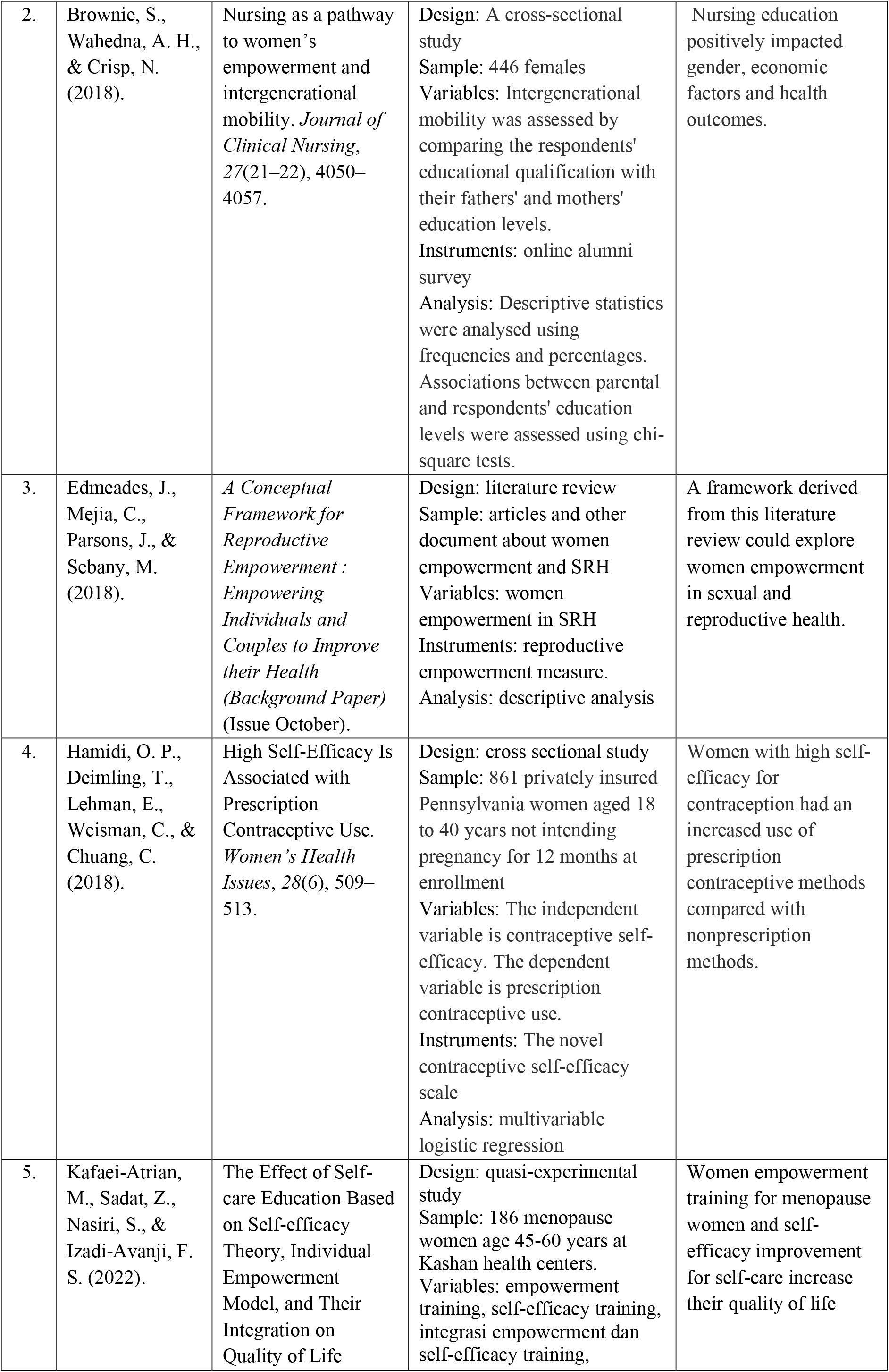
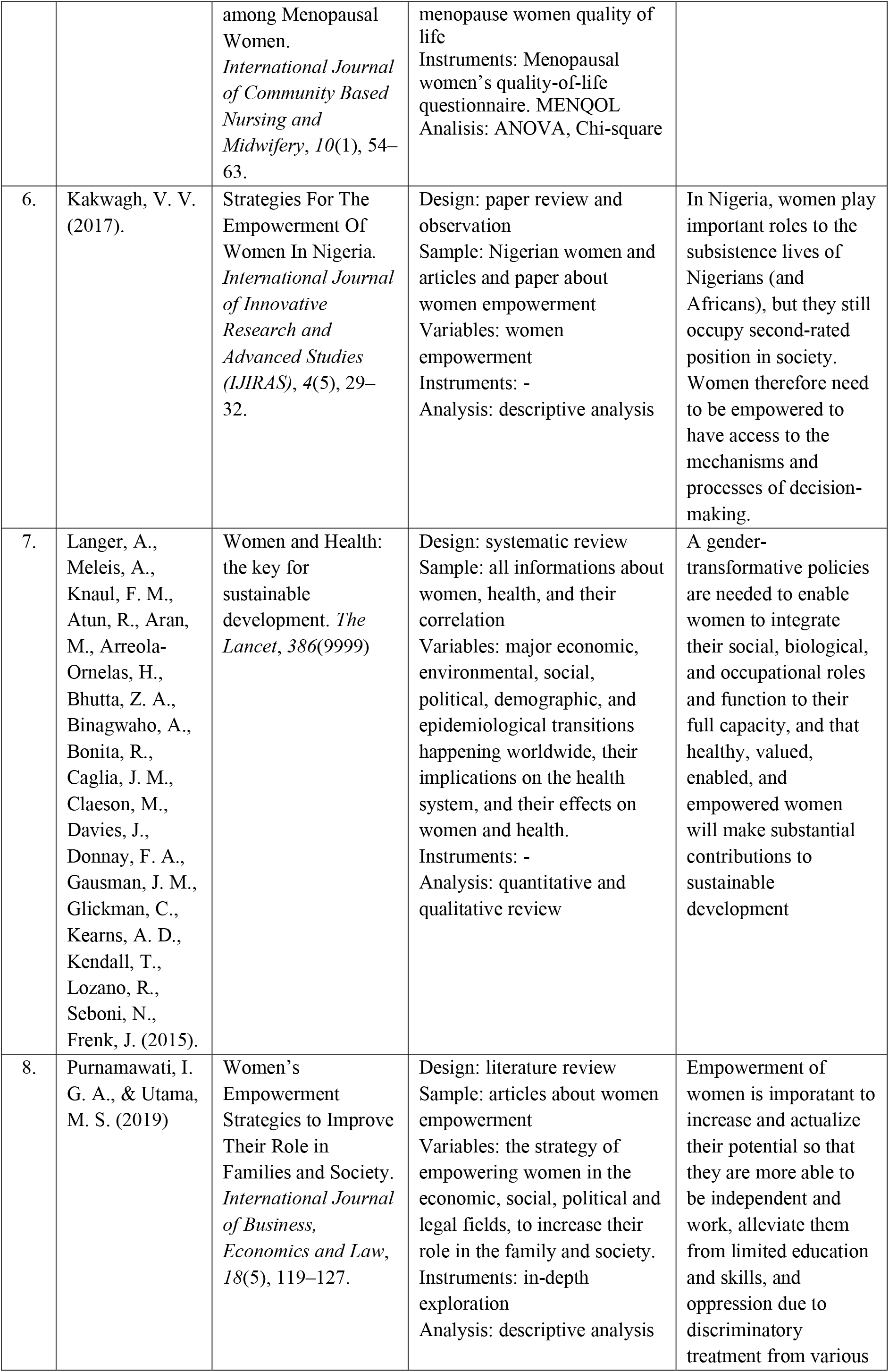
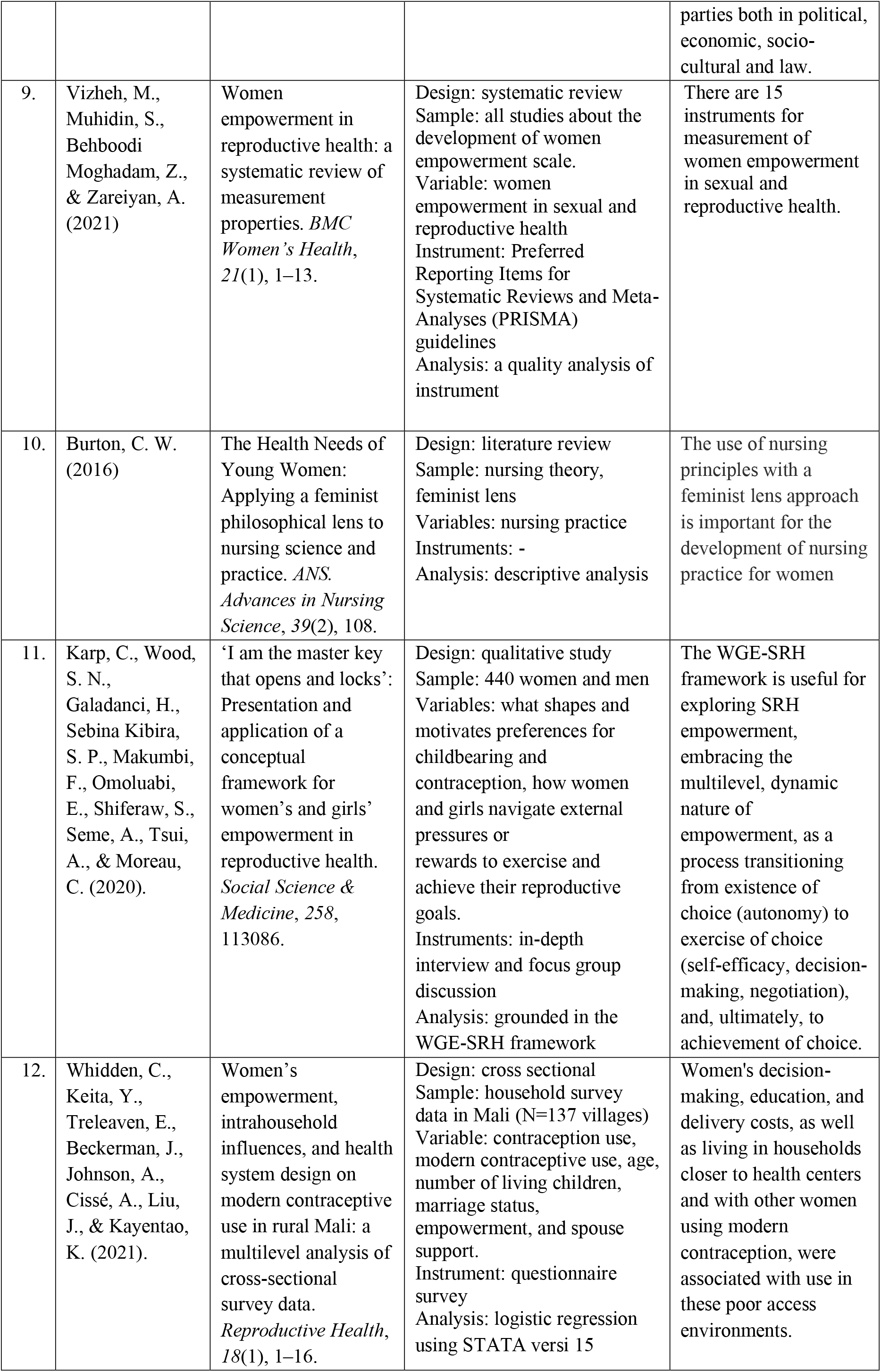
The characteristic of reviewed articles

**Figure 1.**
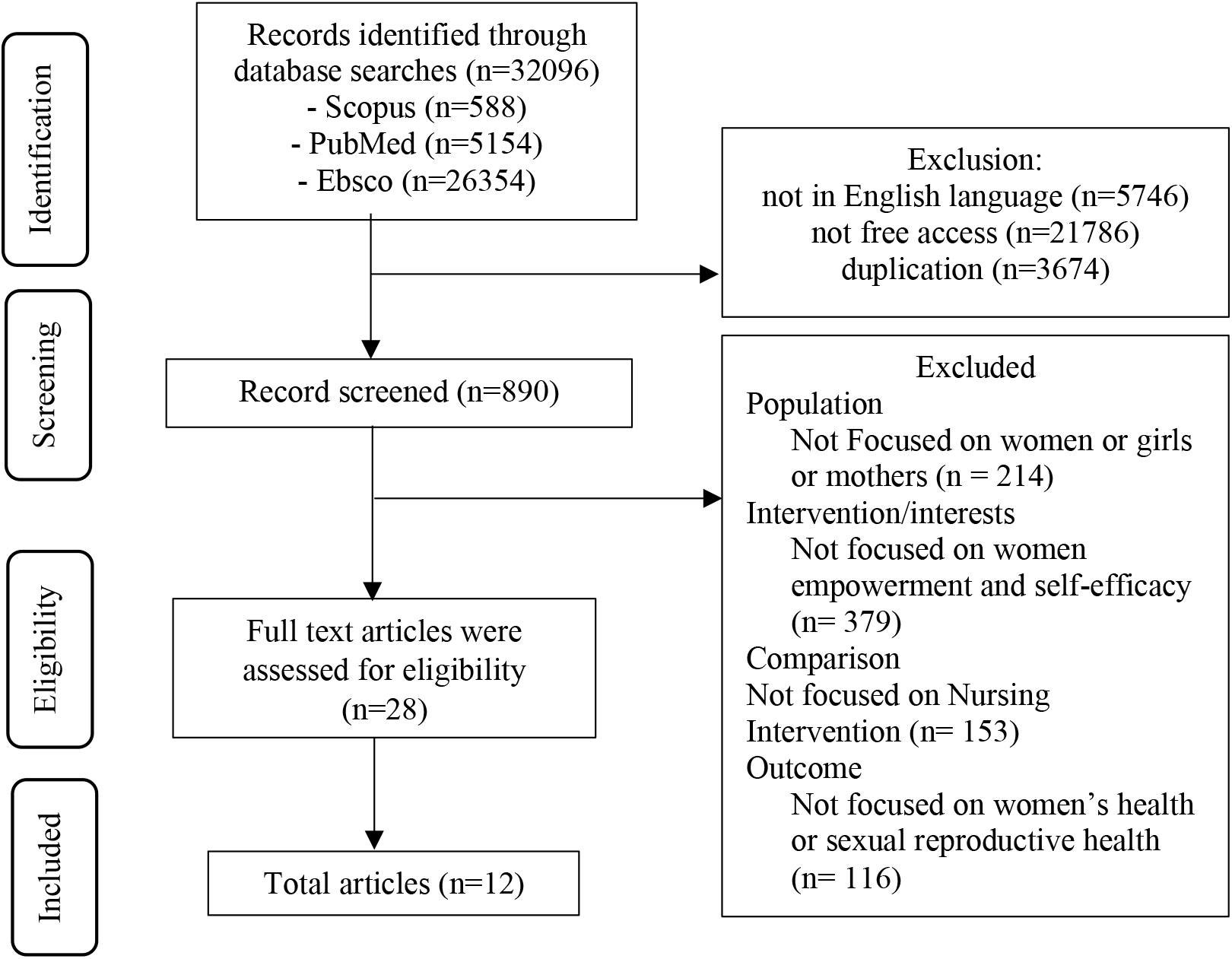
PRISMA Flow Chart.

## Discussion

### Nursing philosophy and women’s health perspective

Women are the pillars of nation-building. Women are the key to the quality of life in a family. Therefore, maintaining women’s health is very important and has become a concern for various parties from year to year (Langer et al., 2015). Nursing as part of health services must be able to comprehensively meet the care needs of women from every stage of life (Burton, 2016). Nursing education has a positive impact on gender because the majority of nurses are women and nursing reflected feminist care. Nurses are also the frontline health provider, therefore, the development of specific nursing care for women’s health needs to be continuously developed (Brownie et al., 2018).

Nursing philosophy is a nurse’s perspective on education, practice, and ethics in carrying out nursing care. The basic principle in the philosophy of nursing is that nursing is a science based on scientific truth. Evidence-based nursing practice is an important approach to developing nursing science through research that can be verified and can be accounted for objectively (Alligood, 2014). The application of nursing philosophy can be done by conducting research, evaluating nursing care feedback, and studying literature to find solutions to problems encountered in the field.

Women have a strategic position in the health sector. The health risks faced by women are different from those of men. This difference is due to differences in the anatomy and physiology that lead to the functions of the reproductive, hormonal, and metabolic systems in the bodies of women and men. The difference is also seen in the roles and responsibilities given by the community to men and women. All of these factors will ultimately affect the way decisions are made, the health risks faced, the efforts made, and how the health system responds to their needs (Matud, 2017). The world’s priorities for women’s health are also constantly evolving. The focus of women’s health, which used to be only on maternal and child health, has now turned wider, namely on aspects of sexual and reproductive health. However, two very basic women’s health problems that need serious attention are the high maternal mortality rate, and the low access to women’s health services (United Nations Population Fund, 2022).

Various previous studies concluded that women’s health is strongly influenced by sociocultural factors that consider women inferior in society. The main factor that becomes the main concern of various parties is ending gender inequality and promoting women’s empowerment to ensure the implementation of human rights. Women must be empowered to maintain their health with a holistic, comprehensive, and lifelong approach. Healthy women will be able to create healthy families and resulting a healthy society and healthy nations (World Health Organization, 2015).

### Concept of Women Empowerment

Empowerment is the ability of individuals or groups to make choices and turn those choices into desired actions and get results from those actions. Empowerment can be interpreted as a process of achieving something such as the ability to make decisions in the family and the ability to plan effectively and is a form of individual control over choices and opportunities in deciding an action. Women’s empowerment is the ability of women to be involved in the decision-making process to achieve equality in social life (Habib Sultan & Yahaya, 2020).

A scientific conceptual framework on women’s empowerment is needed to better understand the concept of women’s empowerment. The conceptual framework that has been widely used to explain women’s empowerment was initiated by Naila Kabeer in 1999 (Vizheh et al., 2021) which explains that women’s empowerment is the ability of women to make choices that are formed from 3 main interrelated dimensions, namely: 1) resources, in the form of material, human, and social; 2) agency, is an individual’s capacity in decision-making, negotiation, manipulation, and resistance; 3) achievement, is the ultimate goal to be achieved by someone. These three dimensions have been widely used as indicators in measuring the level of women’s empowerment in various fields.

### Women empowerment theory in the sexual and reproductive health

Women’s empowerment is one of the strategic steps to improving health status (Edmeades et al., 2018). Empowerment of women in the field of reproductive health is considered important because of the unique nature of the female reproductive system. In addition, sexuality is a fundamental aspect of individual, family, and community life because it is related to reproductive function and is a source of satisfaction to express affection and commitment in a relationship (United Nations Population Fund, 2022). Women with a healthy reproductive system will be able to go through puberty well, avoid infection, go through pregnancy, childbirth, and postpartum healthily and safely, carry out family planning programs well, and can adapt well during menopause.

The conceptual theory of women’s reproductive empowerment initiated in 2016 by Edmeades et.al states that reproductive empowerment is the result of the interaction of 3 factors with multiple processes, namely: 1) voice, showing women’s capacity to carry out their reproductive goals, interests, and desires and have good participation. means in reproductive decision making; 2) choice, which implies the ability of women to make meaningful contributions to reproductive decisions; and 3) power, showing the ability to shape the reproductive decision-making process by exerting power/power over others. Power operates on many levels, including the spouse level, and the family level, and extends to the community and community level (Edmeades et al., 2018).

The framework from Edmeades et al is supported by another framework of Women and Girls Empowerment in Sexual and Reproductive Health (WGE-SRH) which is derived by Karp et.al at Sub-Sahara Africa. The WGE-SRH framework explain that women empowerment started from the existence of choice (autonomy), to exercise of choice (self-efficacy, decisionmaking, negotiation), and to achievement of choice (Karp et al., 2020). Existence of choice corresponds to a woman’s capacity to define her own reproductive goals. Exercise of choice, examines the ability of women and girls to act on their SRH preferences. Achievement of choice is the final step of the empowerment process to achieve the goals. The outcomes of WGE-SRH framework are defined as: pregnancy by choice and contraceptive use by choice (Karp et al., 2020).

Resources are needed to form women empowerment. Relevant resources for individual-level empowerment are comprehensive knowledge, physical health, mental health, and selfefficacy. Comprehensive knowledge about the reproductive system that must be possessed by a woman includes the anatomy and physiology of the reproductive system, fertility, contraception, access to reproductive health services, and women’s rights in reproductive health. This comprehensive knowledge will form a high reproductive health self-efficacy in a woman. There are 4 important components in self-efficacy, namely self-awareness, self-confidence, self-esteem, and the ability to negotiate). Physical and mental health have a significant influence on the formation of individual agency because physically and mentally healthy individuals will be able to implement decisions made related to their reproductive health (Edmeades et al., 2018). WGE-SRH framework also acknowledges the important role of women’s and girls’ resources and opportunity structures, through education, economic conditions, and employment, in shaping women’s and girls’ SRH choices, actions, and achievement of reproductive goals (Karp et al., 2020).

Direct indicators of empowerment must be able to describe the interaction between choice, voice, and power at several different levels and must be flexible to describe empowerment as a process and an outcome. Therefore, the agency which is the center of the empowerment process is expressed in 3 forms of action (Edmeades et al., 2018): 1) Decision making, namely a person’s ability to be actively involved in the process of making a decision. This process is a way to use “voice” and “choice” in each individual’s agency and can occur intrapersonally or interpersonally, 2) Leadership, namely a person’s ability to be the main actor in a decision-making process. make decisions by formulating strategic steps that are beneficial to their welfare through the use of “voice”, “choice”, and “power”, 3) Collective action is a movement carried out by a group of people. to raise status, voice aspirations louder, and increase power, when a goal cannot be achieved by individual action alone. In general, this collective action is used to exert influence on a macrosystem such as at the national or international community level (Edmeades et al., 2018). The conceptual framework allows further development of women’s empowerment models that can be implemented to improve the health status of individuals, families, and communities.

### Strategy to Improve Women’s Empowerment and Self-Efficacy in Sexual and Reproductive Health

Women’s empowerment could be improved by increasing the resources owned by women. The most important resources at the individual level are education and the economy. Education will be able to increase the knowledge, skills, and self-efficacy of a woman so that they can actively participate in various activities in the community. A good education supported by a good economic level will be able to strengthen the empowerment of women. Various studies state that income is one of the causes of women’s low access to health services. Therefore, women must be economically empowered so that they can access the health services needed to maintain the health of themselves and their families. A good family economy is also associated with the ability of the family in terms of fulfilling nutritious food which will also have an impact on the health status of the family (Kakwagh, 2017).

Self-efficacy is the main predictor of women’s empowerment. Self-efficacy refers to confidence in one’s ability to overcome obstacles and achieve desired goals. Previous studies showed that self-efficacy has a strong correlation with women’s empowerment. Research in Saudi Arabia proves that self-efficacy and self-esteem are factors that are strongly related to women’s empowerment among academic and administrative staff at Saudi universities (AlQahtani et al., 2021). A study about contraception concluded that women who have higher selfefficacy are likely more involved in their contraceptive decision-making and choose prescription methods (Hamidi et al., 2018). Another study showed that empowerment and enhancing self-efficacy could improve menopausal women’s quality of life (Kafaei-Atrian et al., 2022). A study on rural Mali showed that women who played any role in decision-making, who had any formal education and participated in any paid labor, were more likely to use modern contraception (Whidden et al., 2021).

Women’s empowerment can occur at the micro-level (empowerment at the individual level by increasing individual capacity), the Meso-level (empowering women in influencing the surrounding environment), and the macro-level (empowering women which allow women to influence policies that have an extensive impact). There are several stages to empowering women, namely: 1) awareness, 2) organizing, 3) cadre regeneration, 4) technical support, and 5) system management. All of these stages will be carried out with the cooperation of various cross-sectoral parties (Purnamawati & Utama, 2019). Such external assistance should be provided by the government as policymakers and the community in the neighborhood where women live. Strengthening government policies should place women’s health services as a priority for health development using cross-sectoral collaboration. While the support of the surrounding community is very important to eliminate the issue of gender inequality to provide opportunities for women to get the same rights as men in terms of education, work, organization, and making decisions related to their health (World Health Organization, 2016).

## Conclusion

Women have enormous potential in nation-building. Women’s health is one of priority in health development. Women’s sexual and reproductive health will affect their general health status. Nursing as part of the health care system has a major role in efforts to improve women’s health status. Through an evidence-based practice approach, it was found that women’s empowerment and women’s self-efficacy are strategic approach for health development. Thus, developing a nursing intervention to improve women’s self-efficacy about sexual and reproductive health could affect women’s empowerment resulting in the improvement of women’s health status.

## Data Availability

All data produced in the present work are contained in the manuscript

